# TCP-25 Gel Reduces Bacterial Burden and Inflammation in Human Epidermal Wounds: Exploratory Analyses from a Randomized Placebo-Controlled Trial

**DOI:** 10.64898/2026.06.26.26356649

**Authors:** Ganna Petruk, Karl Wallblom, Sigrid Lundgren, Bo Nilson, José Cardoso, Ann-Charlotte Strömdahl, Fredrik Forsberg, Congyu Luo, Erik Hartman, Jane Fisher, Karim Saleh, Manoj Puthia, Holger Brüggemann, Artur Schmidtchen

## Abstract

The innate immune system controls bacterial growth and modulates inflammation during wound healing. TCP-25 is a synthetic thrombin-derived host-defense peptide that combines direct antibacterial activity with neutralization of microbial products and modulation of CD14-dependent inflammatory signaling. We investigated whether this dual mechanism translates to human wounds using longitudinal samples from 24 healthy volunteers enrolled in a randomized, double-blind, within-participant, placebo-controlled phase I dose-escalation study of topical TCP-25 gel in matched epidermal suction blister wounds. We assessed inflammatory cytokines, neutrophil-derived proteins, wound exudation, cultivable bacterial burden, spatial bacterial distribution, and microbiome composition. TCP-25 reduced multiple cytokines, myeloperoxidase, and heparin-binding protein, with the strongest effects observed during the peak inflammatory phase. These changes were accompanied by reduced wound exudation and significant reductions in cultivable bacterial burden. Despite this antibacterial effect, microbiome composition and diversity remained largely unchanged, and participant-specific microbial profiles were preserved. TCP-25 therefore coordinated bacterial control, modulation of the physiological inflammatory response, and reduced wound leakage without major disruption of the resident microbiota composition. These findings provide clinical support for translating nature’s endogenous host-defense principles into new therapies for complex wounds.

**Trial registration:** ClinicalTrials.gov NCT05378997

## Introduction

Most skin wounds heal despite continual exposure to microorganisms. However, disruption of the epidermal barrier allows resident and environmental bacteria to access the wound surface, where bacterial growth and microbial products engage pattern-recognition receptors, including CD14 and Toll-like receptors, leading to NF-κB activation and pro-inflammatory cytokine release (Beutler, 2009, Takeuchi and Akira, 2010). Although this early inflammatory response is essential for host defense and repair, excessive bacterial proliferation or sustained activation by pathogen-associated molecular patterns may amplify inflammation, vascular leakage, and wound exudation and thereby interfere with healing (Eming et al., 2007, Gurtner et al., 2008).

Current approaches to bacterial control in wounds rely mainly on antibiotics and antiseptics. These treatments may reduce microbial burden but generally do not directly neutralize the inflammatory effects of bacterial products or modulate the host response that they trigger (Prat et al., 2022, SanMiguel et al., 2018). Broad antimicrobial treatment may also disrupt commensal microbial communities that contribute to skin barrier homeostasis and protection against pathogens. Indeed, clinical studies show that antiseptics such as silver and polyhexamethylenebiguanide may in some cases paradoxically increase infection rates (Saleh et al., 2016, Storm-Versloot et al., 2010). A therapy capable of controlling bacterial expansion while simultaneously moderating microbial-driven inflammation, without disrupting the resident microbiota, would therefore address an important unmet need in wound care.

Wounds have nevertheless healed in microbe-rich environments throughout evolution, suggesting the existence of endogenous mechanisms that coordinate antimicrobial defense with inflammatory control. This reasoning led to the discovery of thrombin-derived C-terminal peptides (TCPs), host-defense peptides released through proteolytic processing of thrombin at sites of tissue injury. TCPs are present in human wound fluids (Saravanan et al., 2017) and combine direct antimicrobial activity with the ability to bind microbial products and modulate innate immune signaling (Papareddy et al., 2010, Puthia et al., 2020, Saravanan et al., 2017).

TCP-25 is a synthetic 25-amino-acid peptide derived from the C-terminal region of human thrombin that encompasses several endogenous TCP sequences. It disrupts bacterial membranes and displays activity against both Gram-positive and Gram-negative bacteria, including *Staphylococcus aureus* and *Pseudomonas aeruginosa* (Papareddy et al., 2010, Puthia et al., 2020). In parallel, TCP-25 binds pro-inflammatory microbial products, including lipopolysaccharide, lipoteichoic acid, and peptidoglycan, and interacts with the microbial ligand-binding site of CD14, thereby reducing downstream Toll-like receptor signaling (Hansen et al., 2015, Saravanan et al., 2018). In infected murine and porcine wounds, topical TCP-25 hydrogel reduced bacterial burden and inflammatory activation and promoted wound healing (Puthia et al., 2020), providing preclinical evidence for a dual antibacterial and immunomodulatory mode of action.

To advance TCP-25 toward treatment of complex wounds, a randomized, double-blind, placebo-controlled phase I study of topical TCP-25 gel in standardized epidermal suction blister wounds in healthy volunteers was conducted (Wallblom et al., 2026). The treatment was safe and well tolerated, with no detectable systemic exposure. Unexpectedly, TCP-25-treated wounds also showed reduced wound leakage (Wallblom et al., 2026), suggesting additional yet unexplored biological effects within the wound environment.

This finding prompted an exploratory investigation of how TCP-25 shapes host-defense responses and bacterial dynamics in human wounds. Using samples from the phase I study, we performed integrated analyses of inflammatory cytokines, neutrophil-derived proteins, wound exudation, cultivable bacterial burden, spatial bacterial distribution, and microbiome composition. We asked whether the dual antibacterial and immunomodulatory activity previously demonstrated in experimental wound models could be reproduced in humans, and whether bacterial control could be achieved while preserving the commensal wound microbiome.

## Results

### Participant demographics

The study design and participant flow are summarized in **Figure 1**. All samples analyzed were obtained from the 24 healthy volunteers enrolled in the parent phase I trial. Full baseline characteristics and concomitant medication have been reported previously (Wallblom et al., 2026). Briefly, the cohort comprised 24 adults (median age 22.5 years, range 18 to 57), including 16 women (67%) and 8 men (33%). Participant characteristics were generally comparable across dose groups, except that all participants in the 2.9 mg/mL-dose group were women. Because each participant received both TCP-25 and placebo in a randomized within-participant design, participant-level baseline characteristics were matched for all treatment comparisons.

**Figure. 1.**
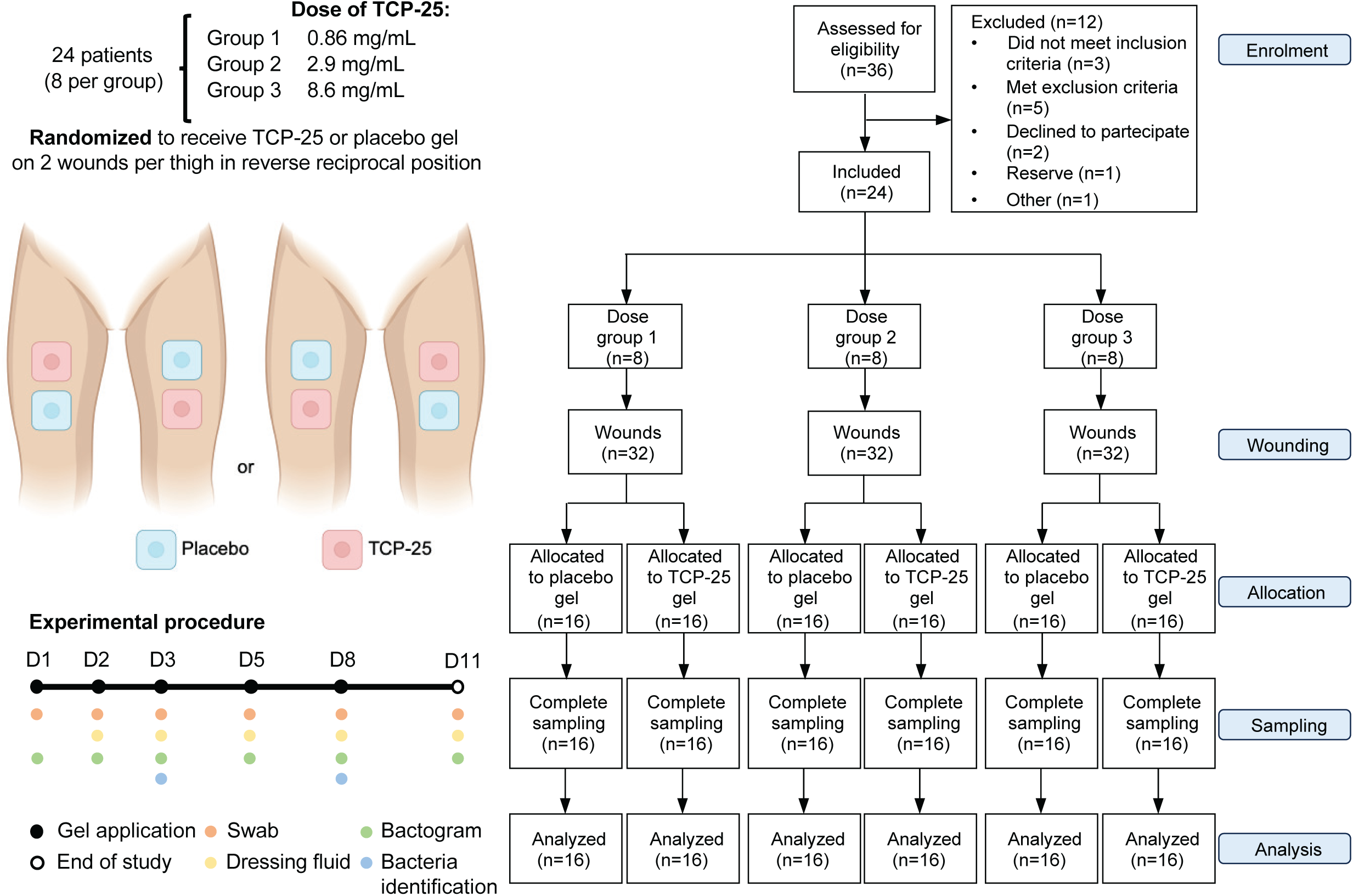
Schematic and CONSORT flow diagram of the randomized, controlled study. This figure was made with BioRender.

### TCP-25 reduces cytokine levels in human wounds

To determine whether the immunomodulatory activity previously observed with TCP-25-containing hydrogel in murine and porcine wound models also occurred in human epidermal wounds, we measured a panel of pro-inflammatory cytokines and chemokines (TNF-α, IL-1β, IL-6, IFN-γ, IL-12p70, and IL-8) and anti-inflammatory or immunoregulatory cytokines (IL-2, IL-4, IL-10, and IL-13) in wound fluids using a V-PLEX assay (**Figure 2a**). Cytokine/chemokine levels in wounds treated with the lowest dose of TCP-25 were similar to those in placebo-treated wounds (**Figure 2b** and **c)**, In the two highest dose groups, the median levels of inflammatory and anti-inflammatory cytokines and chemokines were generally lower, especially on Day 5, in TCP-25-treated wounds versus placebo-treated wounds (**Figure 2b** and **c)**. Median values and confidence intervals for all cytokines per group are found in **Table S1**. Notably, although cytokine/chemokine levels peaked on Day 5 in placebo-treated wounds, they generally remained stable in TCP-25-treated wounds across all time points. Collectively, these findings demonstrate that TCP-25 reduced the levels of both pro- and anti-inflammatory cytokines, suggesting a broad attenuation of immune activation rather than a selective shift toward either a pro- or anti-inflammatory phenotype.

**Figure 2.**
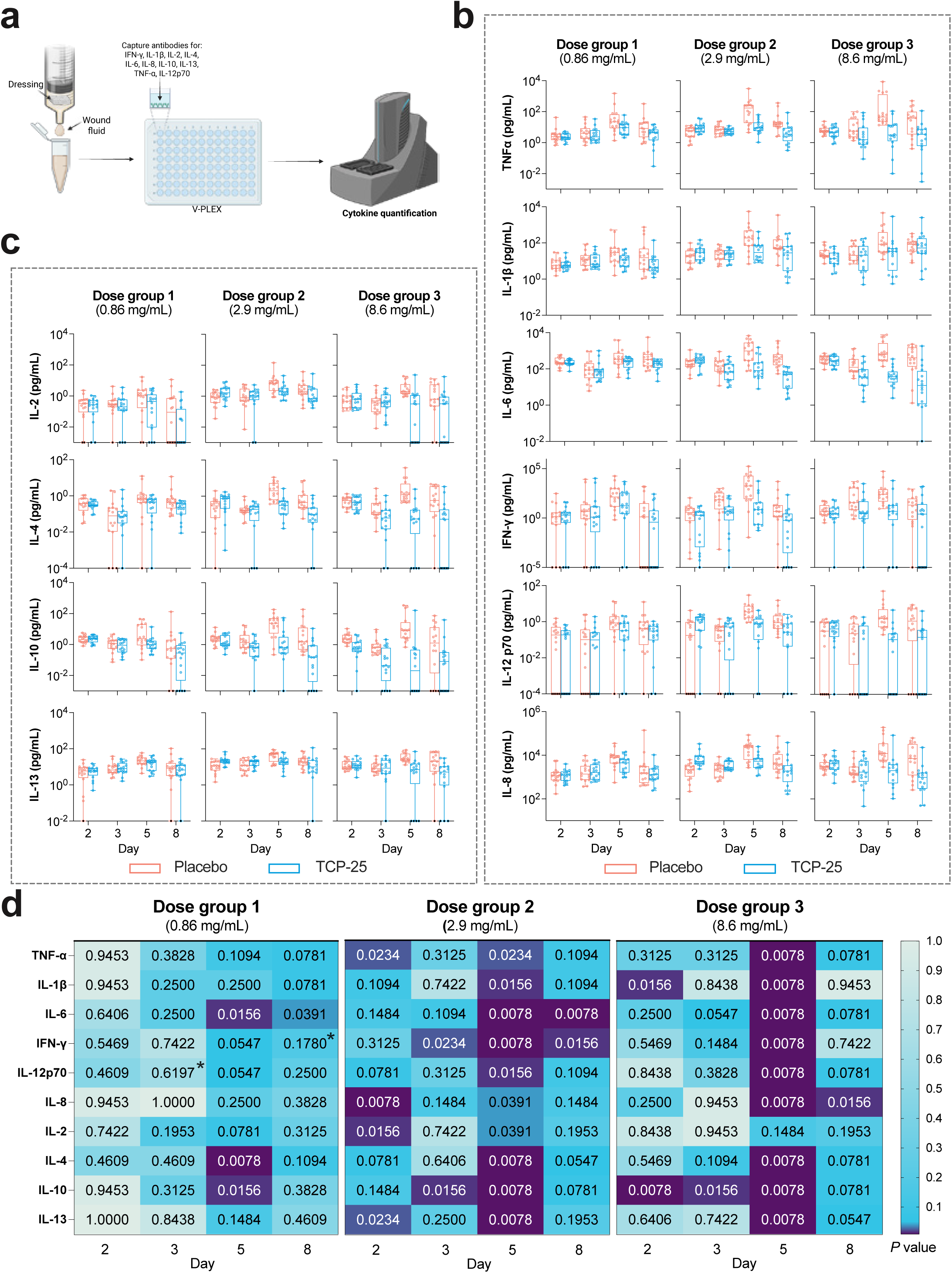
TCP-25-treatment downregulates cytokines in wounds. (**a**) Schematic overview of the wound fluid extract from dressing and subsequent cytokines analysis by V-PLEX. This panel was made with BioRender. **(b, c)** Median levels for each cytokine/chemokine in wounds by dose group over time. The median is represented by a horizontal line, interquartile range by a box, and range by whiskers. Left (L) and right (R) wounds from each participant were plotted separately and treated as individual biological replicates (n=16 wounds per group). Black dots indicate zero values that were replaced to enable plotting on a logarithmic scale. (**d**) Heatmap of *P* values for cytokines and chemokines in wounds treated with TCP-25 compared with placebo-treated wounds. *P* values were calculated using the Wilcoxon matched-pairs signed-rank test. *P* values marked with an asterisk (*) were calculated using the Wilcoxon signed-rank test with Pratt’s method for handling zero differences. For each participant, measurements from the two wounds assigned to the same treatment were averaged, and the resulting mean value was used to calculate the *P* value. *P* values < 0.05 were considered statistically significant.

To identify the cytokines/chemokines and timepoints most affected by TCP-25, we conducted an exploratory post hoc analysis comparing the levels of each mediator in TCP-25-treated versus placebo-treated wounds by paired Wilcoxon test and arranged the resulting *P* values in a heat map (**Figure 2d**). This analysis revealed that the strongest effects of TCP-25 are found on Day 5 in dose group 3. It also identified cytokines that do not fit this pattern, such as IL-2, on which TCP-25 had a weak effect even in the highest dose group, or IL-10, which TCP-25 affected already on Days 2 and 3.

### TCP-25 reduces wound exudation and downregulates neutrophil-associated proteins in wounds

Given that wound exudation is associated with inflammation and neutrophil-derived protein secretion, we examined whether TCP-25 influenced these responses (**Figure 3a**). To quantify fluid accumulation and leakage, the total protein content in wound dressing fluids was measured. Because all dressing fluids were extracted using the same volume of buffer, increased protein concentrations indicate greater wound exudation. At the individual-participant level, TCP-25 reduced wound exudation relative to control, as reflected by decreases in total protein content in dressing fluid, with the strongest effect observed at 8.6 mg/mL (**Figure 3b**). Median values and confidence intervals for total protein content per group are found in **Table S2**. Grouping the individual responses by dose and visualizing them as boxplots revealed a clear dose-dependent pattern, with protein concentrations in dressing fluid from TCP-25-treated wounds remaining lower than placebo from Day 3 onward.

**Figure 3.**
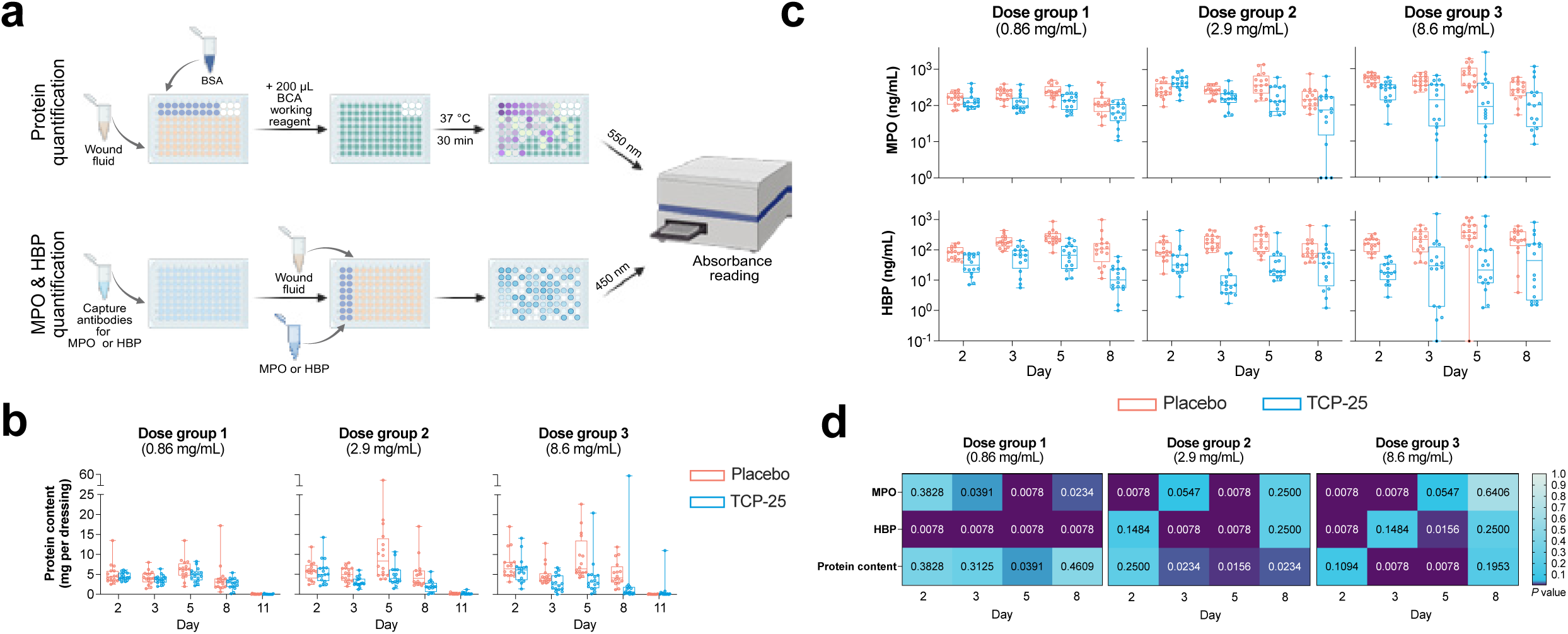
TCP-25-treatment reduces wound exudation and secretion of neutrophil-associated proteins in wounds. **(a)** Schematic overview of the workflow. This panel was made with BioRender. **(b-c)** Levels of protein content per dressing (b), and MPO and HBP levels **(c)** by dose group over time. The median is represented by a horizontal line, interquartile range by a box, and range by whiskers. Left (L) and right (R) wounds from each participant were plotted separately and treated as individual biological replicates (n=16 wounds per group). Black dots indicate zero values that were replaced to enable plotting on a logarithmic scale. (**d**) Heatmap of *P* values for MPO, HBP and protein content in wounds treated with TCP-25 compared with placebo-treated wounds. *P* values were calculated using the Wilcoxon matched-pairs signed-rank test. For each participant, measurements from the two wounds assigned to the same treatment were averaged, and the resulting mean value was used to calculate the *P* value. *P* values < 0.05 were considered statistically significant.

Linear mixed models were constructed to compare protein levels in dressing fluid samples from TCP-25-treated and placebo-treated wounds in each dose group. The models indicate that the protein levels were significantly lower in TCP-25-treated versus placebo-treated wounds on Days 2-8 in each dose group (**Table 1**; *P* = 0.00390, *P* = 0.0001, and *P* = 0.0270 in dose groups 1, 2, and 3, respectively), suggesting that TCP-25 significantly reduces wound exudation on these days.

**Table 1.** Mixed model for protein content in dressing fluid on Days 2, 3, 5, and 8.

| Group | Mean difference in protein content when TCP-25 is used instead of placebo | *P* value |
| --- | --- | --- |
| 1 | -0.93 [-1.55; -0.3] | 0.00390 |
| 2 | -2.93 [-4.34; -1.53] | 0.000100 |
| 3 | -2.26 [-4.27; -0.26] | 0.0270 |

Next, we measured levels of two neutrophil-derived proteins by ELISA in dressing fluid samples (**Figure 3a**). Similar to the patterns that we observed for inflammatory cytokines, median levels of myeloperoxidase (MPO) and heparin-binding protein (HBP) were lower in TCP-25-treated wound versus placebo-treated wounds at most time points and in a dose-dependent manner in all dose groups (**Figure 3c**). Median values and confidence intervals for all neutrophil proteins per group are found in **Table S3**.

To identify the timepoints at which MPO, HBP, and wound exudation were most affected by TCP-25, we conducted an exploratory post hoc analysis comparing their levels in TCP-25-treated versus placebo-treated wounds by paired Wilcoxon test and arranged the resulting *P* values in a heat map (**Figure 3d**). Generally, MPO levels were significantly lower, primarily on Days 2 to 8, for all 3 doses of TCP-25 versus placebo (**Figure 3d**, upper row). Similarly, TCP-25 significantly decreased the levels of HBP across most time points at all doses (**Figure 3d**, middle row). Finally, total protein content was consistently significantly lower in TCP-25-treated wounds on Day 5 in all dose groups, and also on Days 3 and 8 in the higher dose groups (**Figure 3d**, lower row).

### TCP-25 effects persist after normalization to total protein content in wound exudate

To confirm that measured cytokine/chemokine and neutrophil protein levels were not dependent on the total protein content in the dressing fluid, we normalized each variable to the protein content of that corresponding sample (**Figure 4**). Paralleling the results described above, median normalized cytokine/chemokine levels were typically significantly lower in TCP-25-treated wounds on Day 5 in the highest dose groups (**Figure 4a**). Although variability was observed across doses, timepoints, and protein markers, median protein-normalized HBP and MPO levels were significantly lower in TCP-25-treated wounds than in placebo-treated wounds across multiple doses and timepoints (**Figure 4b**). Overall, these results indicate that TCP-25-induced reductions in cytokines/chemokines and neutrophil proteins were not explained by differences in total exudate protein levels.

**Figure 4.**
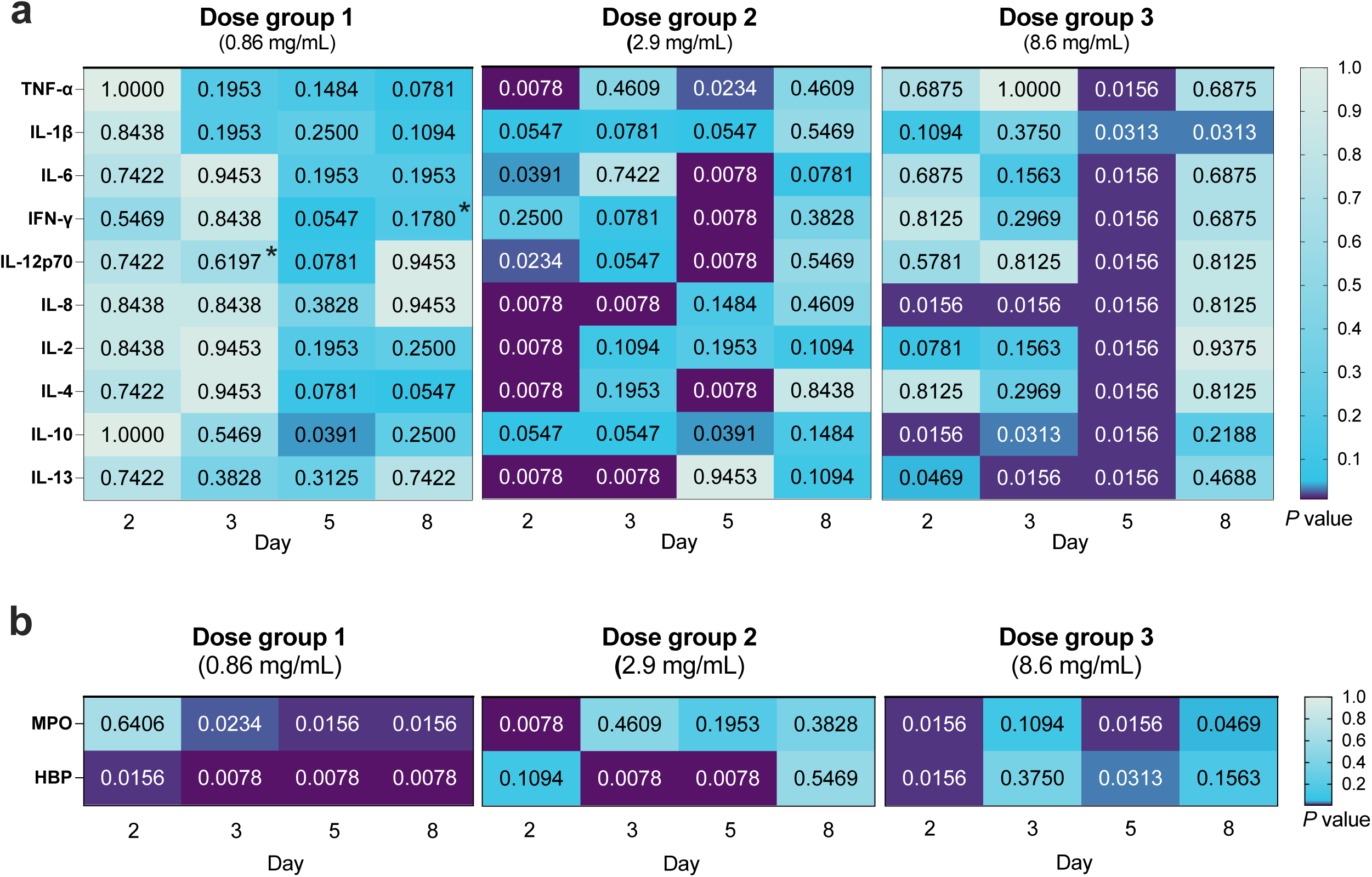
TCP-25 effect on cytokines and neutrophil-related proteins is independent of total protein content in wound exudate. **(a-b)** Heatmap of *P* values for cytokines/chemokines MPO and HBP in wounds treated with TCP-25 compared with placebo-treated wounds. Analyses were performed on cytokines/chemokines, MPO, and HBP quantifications after normalization for protein content. *P* values were calculated using the Wilcoxon matched-pairs signed-rank test. *P* values marked with an asterisk (*) were calculated using the Wilcoxon signed-rank test with Pratt’s method for handling zero differences. For each participant, measurements from the two wounds assigned to the same treatment were averaged, and the resulting mean value was used to calculate the *P* value. *P* values < 0.05 were considered statistically significant.

### TCP-25 dose-dependently decreases aerobic cultivable bacterial levels in wounds

TCP-25 has also been shown to possess antimicrobial activity both in murine and porcine models (Puthia et al., 2020). To verify whether a similar effect occurs in healthy human wounds, we studied aerobic cultivable bacteria in swabs and dressing fluid samples by using different methods (**Figure 5a**).

**Figure 5.**
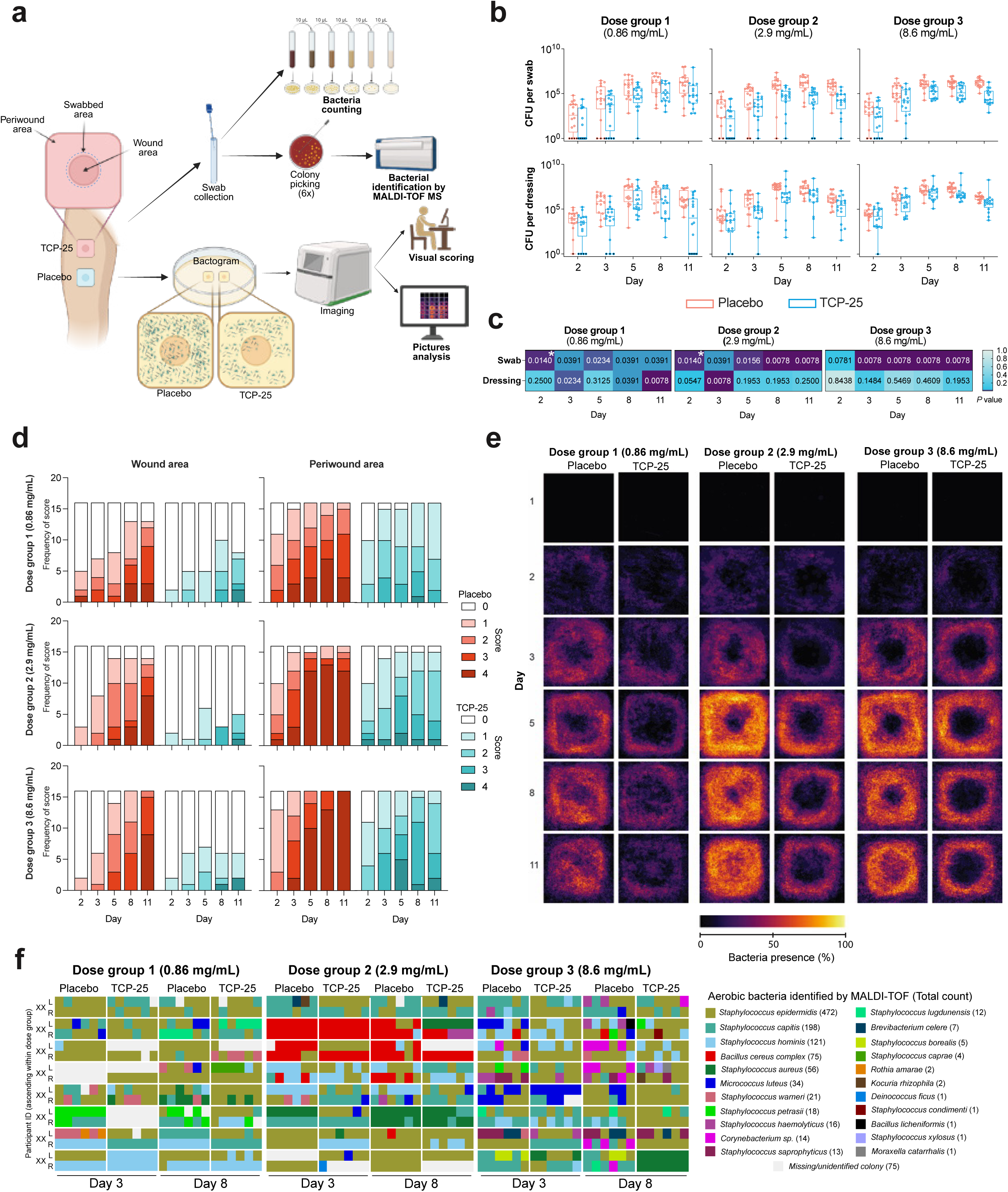
TCP-25 dose-dependently decreases aerobic cultivable bacterial levels in wounds. **(a)** Schematic overview of the workflow. This panel was made with BioRender. **(b)** Boxplots of median bacterial count in swabs (upper panels) and dressings (lower panels) by dose group over time. The median is represented by a horizontal line, interquartile range by a box, and range by whiskers. Left (L) and right (R) wounds from each participant were plotted separately and treated as independent biological replicates (n=16 wounds per group). Black dots indicate zero values that were replaced to enable plotting on a logarithmic scale. **(c)** Heatmap of *P* values for bacterial levels in swabs and dressings from wounds treated with TCP-25 compared with placebo-treated wounds. *P* values were calculated using the Wilcoxon matched-pairs signed-rank test. *P* values marked with an asterisk (*) were calculated using the Wilcoxon signed-rank test with Pratt’s method for handling zero differences. For each participant, measurements from the two wounds assigned to the same treatment were averaged, and the resulting mean value was used to calculate the *P* value. *P* values < 0.05 were considered statistically significant. **(d)** Bacterial coverage levels in the wound and peri-wound area, based on Bactogram method. Bacterial coverage score was assigned by 3 independent raters to each wound. The median number of wounds which were assigned each score is depicted in stacked bar plots. **(e)** Bacterial coverage levels visualized the Bactogram method. Individual Bactogram images from all wounds in each treatment group per day were combined, depicting the frequency with which bacteria were present at each location. Images have been visualized with a colormap to enhance differences. The colormap represents the frequency of bacterial presence at each location as a percentage of the maximum possible frequency (i.e. bacterial presence at that location in all 16 wounds). **(f)** MALDI-TOF colony identification map. For each wound (2 per leg), six bacterial colonies were selected for each wound and identified by MALDI-TOF MS. Each colored square represents a single colony, with colors indicating the bacterial species identified (see legend). Samples are grouped by treatment (placebo vs. TCP-25), dose group (0.86, 2.9, or 8.6 mg/mL), and sampling time point (Day 3 and Day 8). Rows correspond to individual participants (L: left leg, upper row; R: right leg, lower row), and columns correspond to the six colonies analyzed from each sample. White squares indicate no colony identification obtained.

We first determined the colony forming unit (CFU) counts of bacteria per swab and per dressing. Median CFU values and confidence intervals per group are found in **Table S4**. As shown in **Figure 5b** (upper panels), in the swabs, median bacterial levels were lower in TCP-25-treated wounds versus placebo-treated wounds. Notably, these levels remained suppressed in nearly all subjects, all time points, and all 3 doses of TCP-25. Overall bacterial levels followed a similar trajectory in all groups, rising from Days 2-5, plateauing on approximately Day 8, and then declining, except that these levels were lower in TCP-25-treated wounds at each time point. This finding suggests that TCP-25 does not alter the dynamics of bacterial levels in the wound but instead reduces them to similar extents at all time points.

Similarly, median bacterial levels in dressing fluid samples were consistently lower in TCP-25-treated wounds versus placebo wounds over time (**Figure 5b**, lower panels), mirroring its effects in the swabs. These levels had a similar dynamic as the bacterial counts in the swabs, except that they started to decrease earlier, on Day 8.

Linear mixed models were constructed to compare bacterial levels in swabs and dressing fluid samples from TCP-25-treated and placebo-treated wounds in each dose group. Days 1 and 2 were excluded from the mixed models because too many wounds had no bacteria present on those days, and the data were transformed, wherein the dependent variable became the square root of the bacterial count. The models indicate that the bacterial levels in swabs were significantly lower in TCP-25-treated versus placebo-treated wounds over time at each dose (*P* < 0.0001 for all, **Table 2**).

**Table 2.** Mixed model for bacterial levels on Days 3, 5, 8, and 11.

| Sample | Dose Group | Mean difference for sqrt(CFU) (95% confidence interval) when TCP-25 is used | *P* value |
| --- | --- | --- | --- |
| Swab | 1 | -892.26 [-1196.3; -588.26] | <0.0001 |
| Swab | 2 | -815.21 [-1028.5; -601.90] | <0.0001 |
| Swab | 3 | -777.37 [-966.10; -588.65] | <0.0001 |
| DF | 1 | -1311.5 [-1995.9; -627.02] | 0.000200 |
| DF | 2 | -1048.9 [-2459.6; 361.78] | 0.140 |
| DF | 3 | -672.80 [-1908.3; 562.67] | 0.280 |

Sqrt = square root; DF = dressing fluid

The bacterial levels in dressings were also significantly lower in TCP-25-treated wounds compared with placebo-treated wounds over time in wounds receiving 0.86 mg/mL (*P* = 0.0002, **Table 2**), but not in wounds receiving 2.9 or 8.6 mg/mL TCP-25 (*P* = 0.140 and 0.280, respectively, **Table 2**).

To determine the effect size of TCP-25 treatment, the average predicted bacterial loads in the wounds were determined on a population level (**Table 3**). In swabs, TCP-25 treatment reduced bacterial loads by 66.7% to 99.4% on Days 3-8, depending on the dose. In dressing fluid, the magnitude of this reduction was lower.

**Table 3.**
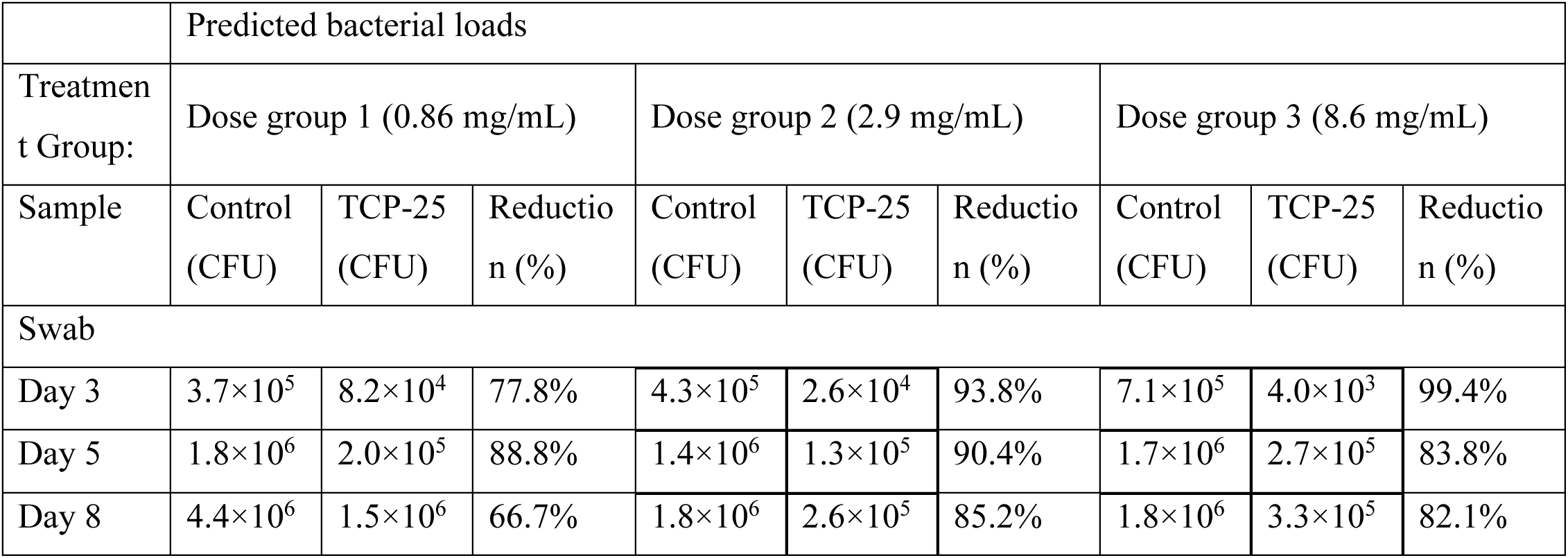
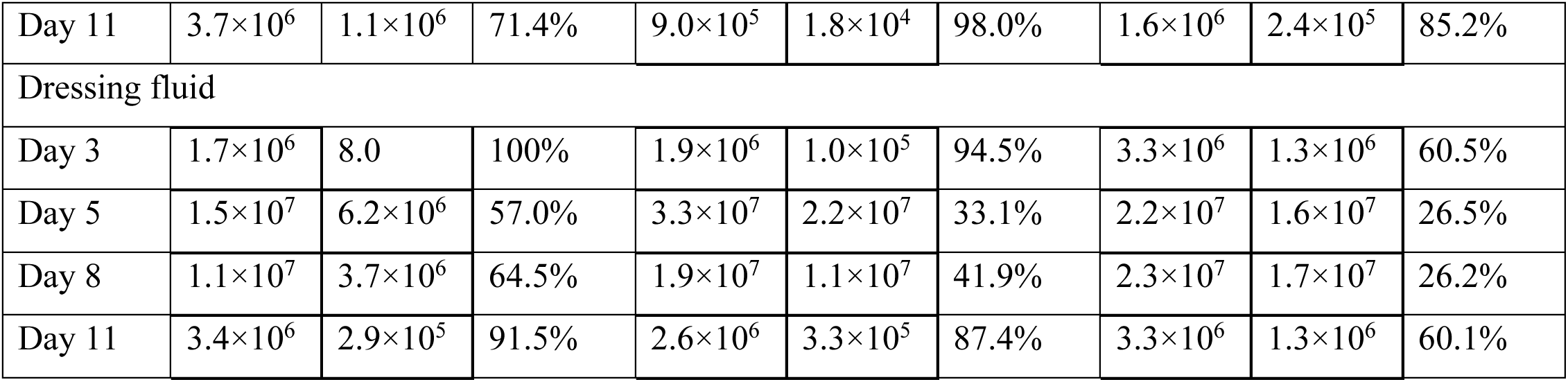
Predicted bacterial loads.

|  | Predicted bacterial loads |  |  |  |  |  |  |  |  |
| --- | --- | --- | --- | --- | --- | --- | --- | --- | --- |
| Treatment Group: | Dose group 1 (0.86 mg/mL) |  |  | Dose group 2 (2.9 mg/mL) |  |  | Dose group 3 (8.6 mg/mL) |  |  |
| Sample | Control (CFU) | TCP-25 (CFU) | Reduction (%) | Control (CFU) | TCP-25 (CFU) | Reduction (%) | Control (CFU) | TCP-25 (CFU) | Reduction (%) |
| Swab |  |  |  |  |  |  |  |  |  |
| Day 3 | 3.7×10<sup>5</sup> | 8.2×10<sup>4</sup> | 77.8% | 4.3×10<sup>5</sup> | 2.6×10<sup>4</sup> | 93.8% | 7.1×10<sup>5</sup> | 4.0×10<sup>3</sup> | 99.4% |
| Day 5 | 1.8×10<sup>6</sup> | 2.0×10<sup>5</sup> | 88.8% | 1.4×10<sup>6</sup> | 1.3×10<sup>5</sup> | 90.4% | 1.7×10<sup>6</sup> | 2.7×10<sup>5</sup> | 83.8% |
| Day 8 | 4.4×10<sup>6</sup> | 1.5×10<sup>6</sup> | 66.7% | 1.8×10<sup>6</sup> | 2.6×10<sup>5</sup> | 85.2% | 1.8×10<sup>6</sup> | 3.3×10<sup>5</sup> | 82.1% |
| Day 11 | $3.7 \times 10^6$ | $1.1 \times 10^6$ | 71.4% | $9.0 \times 10^5$ | $1.8 \times 10^4$ | 98.0% | $1.6 \times 10^6$ | $2.4 \times 10^5$ | 85.2% |
| Dressing fluid |  |  |  |  |  |  |  |  |  |
| Day 3 | $1.7 \times 10^6$ | 8.0 | 100% | $1.9 \times 10^6$ | $1.0 \times 10^5$ | 94.5% | $3.3 \times 10^6$ | $1.3 \times 10^6$ | 60.5% |
| Day 5 | $1.5 \times 10^7$ | $6.2 \times 10^6$ | 57.0% | $3.3 \times 10^7$ | $2.2 \times 10^7$ | 33.1% | $2.2 \times 10^7$ | $1.6 \times 10^7$ | 26.5% |
| Day 8 | $1.1 \times 10^7$ | $3.7 \times 10^6$ | 64.5% | $1.9 \times 10^7$ | $1.1 \times 10^7$ | 41.9% | $2.3 \times 10^7$ | $1.7 \times 10^7$ | 26.2% |
| Day 11 | $3.4 \times 10^6$ | $2.9 \times 10^5$ | 91.5% | $2.6 \times 10^6$ | $3.3 \times 10^5$ | 87.4% | $3.3 \times 10^6$ | $1.3 \times 10^6$ | 60.1% |

To identify the timepoints at which bacterial levels were most affected by TCP-25, we conducted an exploratory post hoc analysis comparing bacterial levels in TCP-25-treated versus placebo-treated wounds by paired Wilcoxon test and arranged the resulting *P* values in a heat map (**Figure 5c**). Overall, bacterial levels in swabs were significantly lower in TCP-25-treated compared with placebo-treated wounds at all timepoints at each dose (upper row). Bacterial levels in dressings were significantly lower in TCP-25-treated wounds compared with placebo-treated wounds at fewer timepoints and only in the lower dose groups (lower row).

The extent of aerobic cultivable bacterial coverage was determined using the Bactogram method (Wallblom et al., 2024). Results for visual scoring and spatial heat map of the wound and periwound areas are shown in **Figure 5d** and **e**, respectively. In the wound area in particular, TCP-25-treated wounds had reduced bacterial coverage, as evidenced by overall lower scores (left panels in **Figure 5d**) and a near-complete absence of bacterial coverage as visualized in the combined Bactogram images (**Figure 5e**). In the periwound area (right panels in **Figure 5d**), TCP-25 reduced but did not completely abrogate bacterial coverage, aligning with the decreased (but not absent) signals around the edge of the dressing area (**Figure 5e**). Thus, indicating that TCP-25 gel provides a local antibacterial effect that is highest in the wound area but extends to the periwound area. The dynamics of bacterial coverage in both areas largely align with the dynamics of bacterial levels measured by quantitative bacterial counts from the swabs and dressings in **Figure 5b**.

Finally, to determine the profile of cultivable aerobic bacteria found in TCP-25-treated and untreated wounds, we performed mass spectrometry analysis. Specifically, we selected 6 colonies per wound on Days 3 and 8, and analyzed them by matrix-assisted laser desorption ionization time-of-flight mass spectrometry (MALDI-TOF MS). The three most frequently detected species belonged to the genus *Staphylococcus*, namely *S. epidermidis*, *S. capitis*, and *S. hominis* (**Figure 5f**). Notably, TCP-25 treatment did not appear to change the overall composition of the cultivable bacteria in the wound at either time point but instead affected their abundance, as indicated by the quantification data (**Figure 5b**).

### Microbiome composition and diversity remain stable during TCP-25 treatment

We assessed the impact of TCP-25 on the wound microbiome by profiling bacterial community composition via 16S rRNA amplicon sequencing at Day 3 and Day 8. At the genus level, samples were heavily dominated by *Staphylococcus* (mean relative abundance 80%, SD 30%), followed by *Bacillus* (6%, SD 22%) and *Corynebacterium* (4%, SD 7.3%). Inter-individual variability appeared to be the primary driver of differences in community composition, with distinct participant-specific profiles often maintained regardless of treatment (**Figure 6a**).

**Figure 6.**
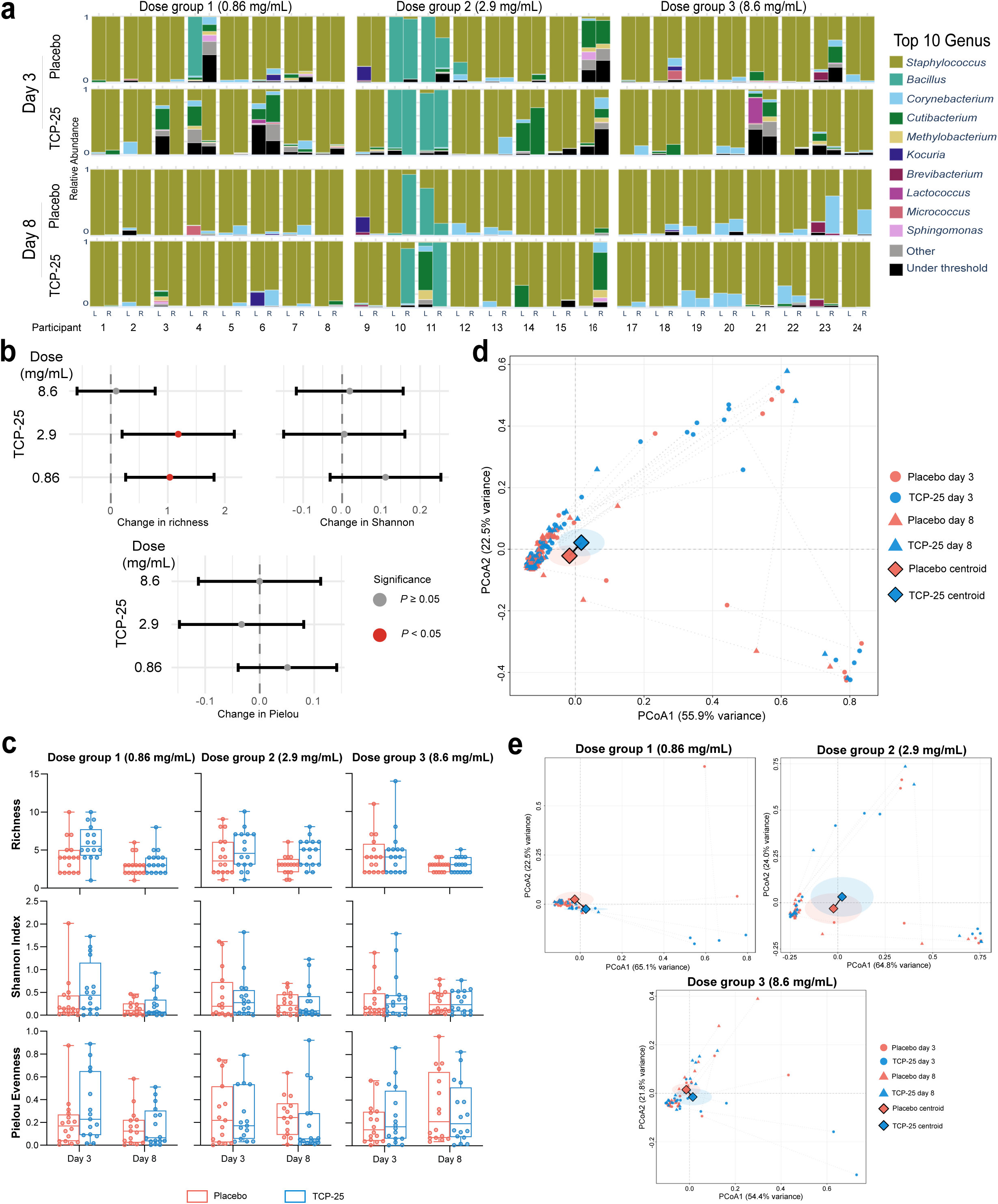
No major shift in microbiome composition at the genus level with TCP-25 treatment compared with placebo treatment. **(a-e)** Genus-level analysis using 16S rRNA (V1–V3) amplicon sequencing. **(a)** Relative abundance of the top 10 bacterial genera. Bars represent individual wound samples (4 wounds per participant) for participants 1–24 at Day 3 and Day 8. **(b)** Forest plots showing the estimated treatment effect (TCP-25 vs. Placebo) on alpha diversity metrics. Points represent estimates from dose-stratified linear mixed models with 95% confidence intervals as lines. Red points indicate statistical significance (*P* < 0.05). Models accounted for longitudinal design and nested wound structure. **(c)** Dose-stratified boxplots of unadjusted alpha-diversity metrics by treatment. For each panel, boxplots display the distribution of richness, Shannon diversity, and Pielou evenness for each TCP-25 dose group at Days 3 and 8, by treatment (TCP-25 vs. placebo). Boxes indicate the interquartile range with the median as a horizontal line; whiskers extend to the minimum and maximum values. Individual points represent wound-level observations (up to 4 wounds per participant). **(b-c)** For richness and Shannon index, n = 8 participants per dose group (64 observations per group). For Pielou evenness, samples with richness ≤ 1 are undefined and thus excluded (2 samples in dose group 1 and 4 samples in dose group 2). **(d-e)** Principal coordinate analysis (PCoA) based on Bray–Curtis dissimilarity including all 192 wound samples (24 participants, all dose groups and time points) **(d)** or for each dose group (8 participants; 64 wounds per group) In **(d–e)**, points are colored by treatment (TCP-25 vs. placebo), with time points indicated by shape. Centroids (diamonds) and 95% confidence interval ellipses represent the mean coordinates for each treatment, either pooled across all dose groups and time points **(d)** or within each dose group, pooled across time points **(e)**. Dotted lines connect paired samples from the same leg (placebo vs. TCP-25) at each time point. A jitter of ±0.015 was applied to improve visualization of overlapping points.

Alpha diversity at the genus level was generally low across all samples, characterized by a median richness of 3 genera (IQR 2 – 5), a Shannon index of 0.05 (IQR 0.05 – 0.46), and a Pielou evenness of 0.16 (IQR 0.05 – 0.34). Overall, linear mixed-effects model analysis indicated that TCP-25 treatment had a negligible impact on alpha diversity compared to placebo (**Figure 6b** and **c**). While TCP-25 treatment was associated with a statistically significant increase in richness at 0.86 and 2.9 mg/mL dose levels, the effect size was modest. The adjusted mean difference in richness was 1.04 (95% CI 0.26 – 1.81; *P* = 0.01) for the 0.86 mg/mL group and 1.18 (95% CI 0.20 – 2.17; *P* = 0.02) for the 2.9 mg/mL group. However, the proportion of variance explained by treatment remained low (partial R² = 0.07 and 0.08, respectively). No significant differences were observed at the highest dose (8.6 mg/mL) for richness, nor for Shannon index or Pielou evenness at any dose level (all *P* ≥ 0.25) (**Table S5**).

Consistent with the taxonomic profiles, Principal Coordinate Analysis (PCoA) based on Bray-Curtis dissimilarity exhibited extensive overlap between the treatment and placebo groups, indicating the absence of large-scale community shifts (**Figure 6d**). This lack of separation was consistent across all dose groups, as illustrated by dose-stratified PCoA plots (**Figure 6e**) and statistically corroborated by permutational multivariate analysis of variance (PERMANOVA) (partial R^2^ ≤ 0.02, *P* ≥ 0.47; **Table S6**). Furthermore, analysis of multivariate dispersion (PERMDISP) indicated homogeneity of group dispersions across treatment arms (*P* ≥ 0.17) (**Table S6**).

Given the dominance of the *Staphylococcus* genus in these wounds, we utilized the *tuf2* amplicon sequencing scheme to achieve species-level resolution. The community was predominantly composed of *S. epidermidis* (62%, SD 37%), followed by *S. capitis* (17%, SD 29%), and *S. hominis* (9%, SD 22%). Consistent with the genus-level findings, visualization of taxonomic profiles indicated that inter-individual variability remained the dominant factor. No consistent shifts were observed between paired TCP-25 and placebo-treated wounds at any dose level (**Figure 7a**).

**Figure 7.**
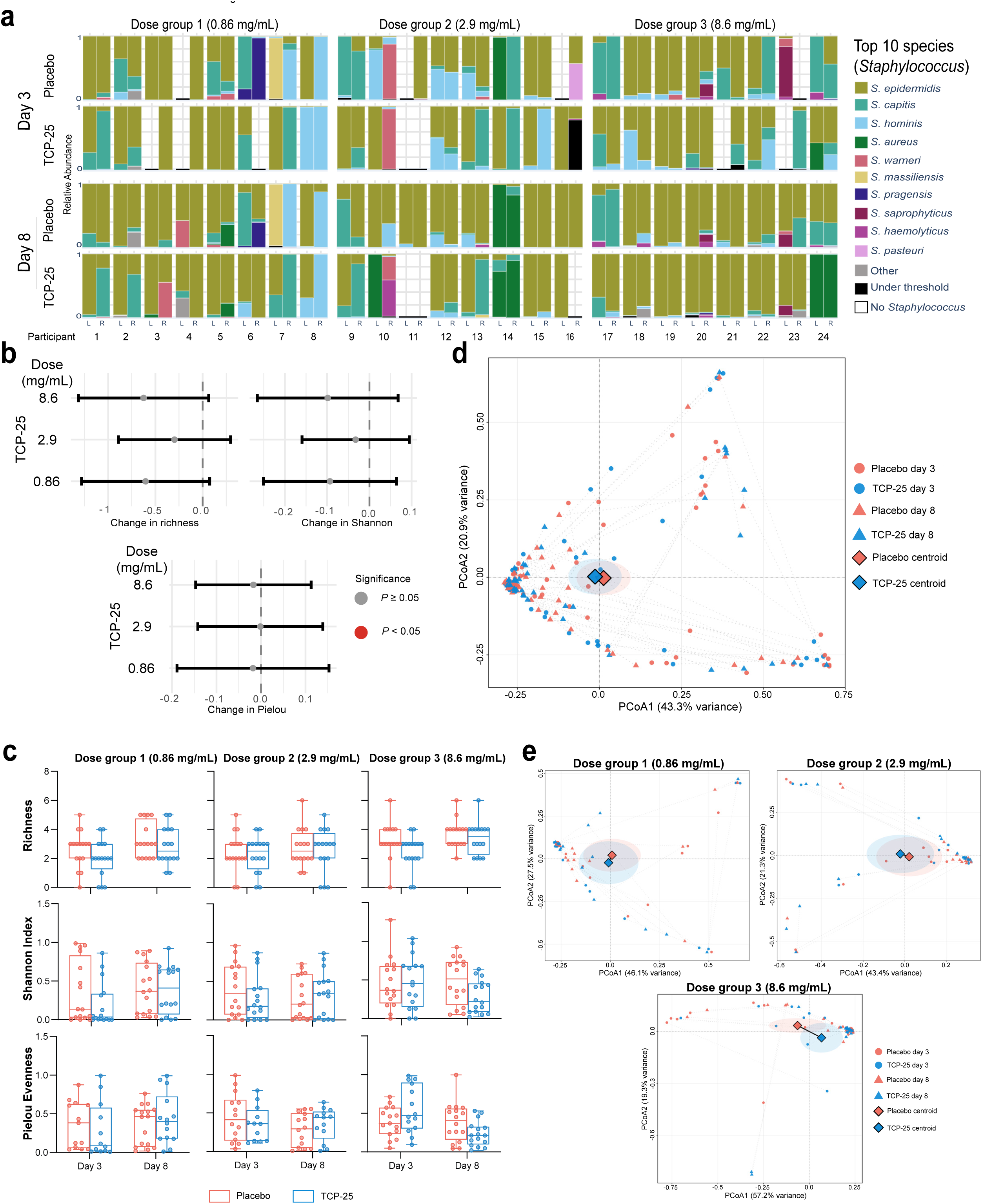
No major shift in microbiome composition at the *Staphylococcus* species level with TCP-25 treatment compared with placebo treatment. (a-e) *Staphylococcus* species-level analysis using *tuf2* amplicon sequencing. **(a)** Relative abundance of the top 10 *Staphylococcus* species. Empty bars indicate samples with no quantifiable staphylococci. **(b)** Forest plots showing the estimated treatment effect (TCP-25 vs. Placebo) on alpha diversity metrics within the *Staphylococcus* genus. Points represent estimates from dose-stratified linear mixed models with 95% confidence intervals as lines. Red points indicate statistical significance (*P* < 0.05). Models accounted for longitudinal design and nested wound structure. **(c)** Dose-stratified boxplots of unadjusted alpha-diversity metrics by treatment. For each panel, boxplots display the distribution of richness, Shannon diversity, and Pielou evenness for each TCP-25 dose group at Days 3 and 8, by treatment (TCP-25 vs. placebo). Boxes indicate the interquartile range with the median as a horizontal line; whiskers extend to the minimum and maximum values. Individual points represent wound-level observations (up to 4 wounds per participant). **(b-c)** Samples with zero staphylococcal abundance were included for richness and Shannon (n = 8, 64 observations per dose group). For Pielou evenness, samples with richness ≤ 1 were excluded. Dose group 1, n = 8 (55 observations); dose group 2, n = 7 (50 observations); dose group 3, n = 8 (61 observations). **(d-e)** Principal coordinate analysis (PCoA) of *Staphylococcus* species based on Bray–Curtis dissimilarity for all three dose groups **(d)** or for each dose group separately **(e)**. Samples with zero staphylococcal abundance were excluded as dissimilarity is undefined for empty vectors. Participant 11 was excluded as all TCP-25 treated wounds had zero staphylococcal abundance. The plot **(d)** includes 175 samples from 23 participants across all dose groups. The plots **(e)** include dose group 1, 8 participants (60 observations); dose group 2, 7 participants (54 observations); dose group 3, 8 participants (61 observations). In **(d–e)**, points are colored by treatment (TCP-25 vs. placebo), with time points indicated by shape. Centroids (diamonds) and 95% confidence interval ellipses represent the mean coordinates for each treatment, either pooled across all dose groups and time points **(d)** or within each dose group, pooled across time points **(e)**. Dotted lines connect paired samples from the same leg (placebo vs. TCP-25) at each time point. A jitter of ±0.015 was applied to improve visualization of overlapping points.

Species-level alpha diversity within the *Staphylococcus* genus was characterized by a median richness of 3 species (IQR 2 – 4), a Shannon index of 0.36 (IQR 0.05 – 0.64), and a Pielou Evenness of 0.37 (IQR 0.13 – 0.55). As illustrated in the forest plot (**Figure 7b**), model estimates for TCP-25 treatment effects were small across all dose groups and alpha metrics. Consistent with this, no statistically significant differences were observed for richness, Shannon index, or Pielou evenness at any dose level (all *P* ≥ 0.15), and the estimated proportion of variance explained by treatment remained low (partial R^2^ ≤ 0.04) (**Table S7**). Boxplots of alpha diversity scores per treatment and dose group are presented in **Figure 7c**.

Visualization of beta diversity using PCoA plots based on Bray-Curtis dissimilarity showed greater inter-sample variability at the species level than at the genus level, yet no clear clustering by treatment arm was evident (**Figure 7d**). While dose-stratified PERMANOVA detected a small but statistically significant compositional shift in the highest dose group (8.6 mg/mL) after adjustment for temporal effects (partial R^2^ = 0.04, *P* = 0.04), this finding was not reflected by distinct clustering in the corresponding PCoA plot (**Table S8, Figure 7e**). No significant compositional shifts were observed in the lower-dose groups (0.86 and 2.9 mg/mL), ≤1% of the variance (*P* = 1.0). Furthermore, PERMDISP analysis indicated homogeneity of group dispersions across treatment arms (*P* ≥ 0.19) (**Table S8**).

Taken together, the 16S rRNA and *tuf2* sequencing analyses showed that TCP-25 treatment did not induce broad changes in wound microbiome composition or diversity. Instead, microbial profiles were largely shaped by inter-individual differences, with participant-specific communities maintained despite reduced cultivable bacterial levels.

## Discussion

In this exploratory analysis of a randomized first-in-human study, topical TCP-25 reduced inflammatory cytokines, neutrophil-associated proteins, wound exudation, and cultivable bacterial levels in acute epidermal wounds. Importantly, these effects occurred without major changes in the composition or diversity of the wound microbiome. The findings extend the previously demonstrated dual antibacterial and immunomodulatory activity of TCP-25 from experimental wound models (Puthia et al., 2020) to human wounds.

Inflammation is essential during the early phase of wound healing and is required for microbial control, clearance of damaged tissue, and initiation of repair. TCP-25 reduced both pro-inflammatory and immunoregulatory cytokines, with the strongest effects observed around Day 5, when cytokine levels peaked in placebo-treated wounds. This broad effect indicates a general attenuation of immune activation rather than selective modulation of individual cytokine pathways. Importantly, cytokine responses remained detectable, showing that TCP-25 modulated rather than abolished the physiological inflammatory response.

These findings agree with the known mechanism of TCP-25. The peptide binds bacterial products, including LPS, LTA, and peptidoglycan, and interacts with the microbial ligand-binding pocket of CD14, thereby reducing downstream TLR-mediated signaling (Hansen et al., 2015, Saravanan et al., 2018). Previous studies demonstrated that these effects reduce NF-κB activation and cytokine responses in vitro and in experimental wound infection models (Puthia et al., 2020, Stromdahl et al., 2021). The present clinical data therefore support the concept that TCP-25 limits amplification of inflammation triggered by bacteria and their products without completely inhibiting the physiological inflammatory response during wound healing.

It could be argued that the reduced inflammatory responses were secondary to the antibacterial effect of TCP-25, because lower bacterial levels would also reduce the amount of pro-inflammatory microbial products in the wound. This indirect mechanism likely contributes to the overall response but does not fully explain the findings. All three TCP-25 doses significantly reduced cultivable bacterial levels, whereas the broad and most consistent reductions in cytokines were mainly observed at the two higher doses and during the peak inflammatory phase. Moreover, a separate detailed analysis of the placebo-treated wounds from the same phase I study showed that bacterial levels correlated only weakly with most cytokines and showed moderate associations mainly with TNF-α, IL-1β, and IL-10 around Day 5, while correlations with neutrophil proteins and wound exudation were minimal or absent (Lundgren et al., 2025). These observations suggest that bacterial abundance is only one determinant of wound inflammation. Together with previous studies demonstrating direct neutralization of microbial products and inhibition of CD14-dependent signaling by TCP-25 (Puthia et al., 2020, Saravanan et al., 2018), the present data support a model in which antibacterial and direct immunomodulatory effects act in parallel.

TCP-25 also reduced the neutrophil-derived proteins MPO and HBP, which contribute to antimicrobial defense, inflammatory signaling, vascular responses, and leukocyte recruitment (Odobasic et al., 2013, Soehnlein and Lindbom, 2009, Young et al., 2004), together with IL-8, a major neutrophil-recruiting chemokine (Ellis et al., 2018). These findings indicate reduced neutrophil-associated activation in TCP-25-treated wounds. Because bacterial levels showed minimal or no correlation with neutrophil proteins in the placebo-treated wounds (Lundgren et al., 2025), the reductions in MPO and HBP are unlikely to be explained primarily by lower bacterial burden. They may instead reflect attenuation of microbial product-induced signaling and direct modulation of neutrophil activation and migration, as previously demonstrated for TCP-25 (Lim et al., 2017). However, neutrophil numbers, cellular sources of cytokines, and leukocyte trafficking were not directly assessed in the present study, and the relative contribution of these mechanisms remains to be determined.

A particularly notable effect was the reduction in wound exudation. This initial clinical observation (Wallblom et al., 2026) was supported by lower total protein levels in dressing fluids, with the clearest effects observed during Days 3–8. Wound exudate is required for normal healing, but excessive leakage may cause maceration, increase dressing requirements, and complicate the management of chronic wounds (Walker and Brace, 2019). Of note is that TCP-25 reduced rather than abolished exudation, suggesting a modulatory effect on fluid leakage during the inflammatory phase. The underlying mechanism was not directly studied, but the reduction in HBP provides a possible explanation. HBP is released from activated neutrophils and is a potent inducer of vascular permeability (Soehnlein and Lindbom, 2009). It is also elevated in wound fluids from chronic non-healing wounds (Lundqvist et al., 2004). Thus, reduced neutrophil activation and HBP release may contribute to the decreased wound leakage observed after TCP-25 treatment. This hypothesis will require direct studies of vascular permeability and fluid transport.

The antibacterial effect of TCP-25 was consistently observed in wound swabs across all three dose groups. Depending on dose and time point, predicted bacterial levels were reduced by approximately 67–99% compared with placebo-treated wounds. Notably, the temporal development of bacterial colonization was similar in treated and control wounds, but bacterial levels remained lower after TCP-25 treatment. TCP-25 therefore reduced bacterial abundance without disrupting the normal dynamics of colonization in this acute wound model.

An important finding was that this reduction in cultivable bacteria was not accompanied by a major shift in microbiome composition. The wounds were dominated by commensal staphylococci, particularly *S. epidermidis*, *S. capitis*, and *S. hominis*. Inter-individual differences were the main determinant of microbiome composition, whereas TCP-25 treatment explained only a small proportion of the observed variation. Alpha diversity remained largely unchanged, and no clear treatment-related clustering was observed in the beta-diversity analyses.

These results indicate that TCP-25 reduced viable bacterial levels without broadly altering the composition of the resident wound microbiota. This distinction is important. Commensal skin bacteria contribute to skin barrier homeostasis, colonization resistance, and regulation of immune responses (Canchy et al., 2023, Tomic-Canic et al., 2020, Zheng et al., 2022). A treatment that controls bacterial expansion while preserving the overall composition of the commensal community may therefore offer advantages over broad antimicrobial approaches that can disturb microbial ecology.

Taken together, the reduction in bacterial burden, cytokines, neutrophil-associated proteins, and wound exudation supports the proposed multimodal mechanism of TCP-25. Direct antibacterial activity lowers viable bacterial levels, whereas binding of bacterial products and modulation of CD14-dependent signaling limit amplification of the inflammatory response. These mechanisms are likely to act in parallel and reinforce each other. Lower bacterial abundance reduces the amount of pro-inflammatory microbial material, while neutralization of bacterial products reduces the host response to bacteria that remain in the wound.

This combined mechanism differs from that of conventional antibiotics and antiseptics, which are primarily designed to kill or inhibit microorganisms and do not directly target the inflammatory activity of microbial products. TCP-25 is derived from an endogenous wound-defense system in which antimicrobial activity and regulation of innate immunity are integrated within the same molecule. The present findings provide clinical support for this concept.

Importantly, TCP-25 has subsequently also been evaluated in patients with dystrophic epidermolysis bullosa (EB), where topical treatment was safe and positive exploratory signals for wound healing were observed (Wallblom et al., 2025). Although that study was not designed to establish efficacy, it demonstrated that TCP-25 can be applied safely to fragile and chronically wounded skin. Together with the present findings, these clinical data support further development of TCP-25 for complex wounds characterized by bacterial colonization, inflammation, exudation, and impaired healing.

The study has several strengths. The standardized suction blister model enabled controlled comparisons in acute human wounds, and the reciprocal within-participant design reduced the influence of inter-individual differences in inflammatory and microbial responses. This was particularly important given the marked subject-specific microbiome profiles. Furthermore, the use of several complementary methods allowed the effects of TCP-25 to be assessed at multiple levels, including cytokines, neutrophil proteins, wound exudation, cultivable bacterial burden, spatial bacterial distribution, bacterial identification, and microbiome composition.

The study also has limitations. It included only 24 healthy volunteers and was designed primarily to assess safety, tolerability, and pharmacokinetics (Wallblom et al., 2026). The present analyses were exploratory, were not prospectively powered for efficacy endpoints, and included multiple comparisons. The findings should therefore be considered hypothesis-generating and require confirmation in larger studies. In addition, the wounds were acute and normally healing and did not reproduce the complex inflammatory, microbial, vascular, and tissue environments of infected or chronic wounds. The results cannot therefore be directly extrapolated to chronic ulcers, burns, infected wounds, or wounds in EB. Finally, vascular permeability was not directly measured.

In conclusion, topical TCP-25 reduced bacterial burden, inflammatory activation, neutrophil-associated proteins, and wound leakage in acute human wounds without major disruption of the commensal microbiome composition. These findings support further evaluation of TCP-25 in wounds characterized by bacterial overgrowth, inflammation, and excessive exudation. More broadly, they show how principles evolved by nature for millions of years to coordinate microbial control and inflammation can be translated into new wound therapeutics.

## Methods

### Study design

Samples analyzed in this exploratory study were obtained from participants enrolled in a randomized first-in-human study (clinicaltrials.gov identifier NCT05378997). Details of the parent trial design and methodology have been published previously (Lundgren et al., 2023). No changes relevant to the present exploratory analyses were made after trial commencement.

### Treatment

The 24 healthy adults included in the study were divided into 3 groups (8 subjects each) (**Figure 1**). Each subject received 4 similarly sized suction blisters in total (2 on each thigh). The 3 groups were assigned to receive a topical dose of TCP-25 gel—0.86, 2.9, or 8.6 mg/mL. Within each dose group, each subject was administered TCP-25 gel and placebo on 1 wound each on the same thigh in a randomized manner; the wounds on the other thigh were treated similarly but in the reverse reciprocal position.

The respective doses were administered on Days 1, 2, 3, 5, and 8. Bactograms, swab, and dressing fluid samples were collected at the designated time points, as shown in **Figure 1**.

### Swab procedure

Swabbing was performed as described by (Lundgren et al., 2025). Briefly, each wound was swabbed on Day 1, 2, 3, 5, 8, and 11 using a sterile cotton swab (Selefa, Stockholm, Sweden) pre-wetted with sterile phosphate-buffered saline (PBS). Then the swab was placed into a microfuge tube with 0.5 mL sterile PBS and kept on ice until analyses (no later than 2 hours after sampling).

### Dressing extraction

Mepilex dressing (Mölnlycke Healthcare, Gothenburg, Sweden), used to cover the wounds in between study visits, were changed on Days 2, 3, 5, 8, and 11. The wound exudate were extracted as described by (Lundgren et al., 2025). Briefly, the Mepilex dressing was placed into an empty 5 mL conical tube and kept on ice before weighing it in the tube. After, the dressing was placed into a 5 mL syringe containing 2 mL of cold, sterile 10 mM Tris buffer, pH 7.4. The syringe was vortexed for 5 minutes and the fluid was extracted by depressing the plunger and emptying the contents into the original collection tube. The extracted fluid was kept on ice until being used for quantification of bacterial levels. The leftover volume was divided in 2 aliquots, one was aliquoted and frozen at –80°C as it is, the other one was first complemented with 100× Halt Protease Inhibitor Cocktail (Thermo Fisher Scientific, USA) to a final strength of 1×, and then aliquoted and frozen at –80°C. Protein content, neutrophil proteins, and cytokines in the dressing fluid samples that contained protease inhibitors were quantified, as described below.

### Quantification of protein content

Protein content in dressing fluids was calculated by bicinchoninic acid (BCA) assay (Thermo Fisher, Waltham, MA, USA), following the manufacturer’s instructions. The concentration of protein in mg/mL was calculated from the standard curve obtained by using bovine serum albumin. The total protein content was determined by multiplying it to the total volume of dressing fluid extract, calculated as described by (Lundgren et al., 2025). The assessor was not blinded to the identity of the assigned intervention for this measure.

### Quantification of cytokines and neutrophil proteins

Cytokines (interferon gamma [IFN]-γ, interleukin [IL]-1β, IL-2, IL-4, IL-6, IL-8, IL-10, IL-12p70, IL-13, and tumor necrosis factor [TNF]-α) were measured as described by (Lundgren et al., 2025). Briefly, V-PLEX Proinflammatory Panel 1 (human) kit (Meso Scale Diagnostics, art.no: K15049D) was used following the manufacturer’s instructions. Each sample was tested at 5× dilution. Samples for which IL-8 was above the upper limit of detection were analyzed again using the U-PLEX Biomarker Group 1 (human) kit (Meso Scale Diagnostics, art.no: K151TYK-2) at a higher dilution. Cytokine concentration in each sample was calculated using the MSD discovery Workbench analysis software. Samples with a concentration below the detection limit were not quantified by the analysis software and were assigned a value of zero for all analyses. The number of samples (N = 32) with cytokine concentrations falling within the range of the standard curve for each cytokine is summarized in **Table S9**.

Human myeloperoxidase (MPO) and heparin-binding protein (HBP) were measured using ELISA kits from R&D Systems (Minneapolis, MN, USA) and Aviva Systems Biology (San Diego, CA, USA), respectively. Samples were analyzed in duplicate following the manufacturer’s instructions. Absorbance was read at 450 nm on a microplate reader (BioRad, Hercules, CA, USA). A standard curve was established with reference standards to calculate the concentration in each sample.

For all measurements of neutrophil proteins and cytokines, the assessor was not blinded to the identity of the assigned intervention.

In some analyses, to obtain the amount of each cytokine or neutrophil protein levels per mg of protein in the dressing fluid extract sample, cytokine and neutrophil protein levels were normalized for the protein concentration measured by BCA assay, as described above.

### Quantitative bacterial counts

To quantify the viable cultivable bacteria, the swabs and dressing fluid samples were analyzed as described previously by (Lundgren et al., 2025). Briefly, samples were diluted with sterile PBS to generate 7 10-fold serial dilutions (from 10× to 10^7^×). Then, six separate 10-μL drops of the undiluted sample and each of the dilutions were deposited on a Todd-Hewitt (TH) agar plate. After drying, the plates were incubated at 37°C in 5% CO_2_ overnight. The next morning, the number of colonies was counted and colony forming units (CFU)/mL was calculated. Then, obtained CFU/mL was multiplied by the total volume of fluid in the sample (0.5 mL for swabs or the volume calculated from the dressing weight described above for dressing fluid extract) to obtain the total CFU per swab and CFU per dressing. Assessors were blinded to the identity of the assigned intervention for this measure.

### Bactogram procedure

Spatial mapping of culturable aerobic surface bacteria was performed using the Bactogram method as described previously (Wallblom et al., 2024). Briefly, sterile (autoclaved) 12.5 cm filter paper (Grade 3, Ahlstrom-Munksjo, Sweden) was pre-wet with 2 mL sterile PBS before being transferred to the wound. The filter paper was positioned to cover both wounds on each leg and, using the lid of a petri dish, was pressed gently on the wounds for 1 minute with a rocking motion to ensure equal contact of the wounds with the filter paper. The filter paper was then placed face down on a 15 cm agar plate and incubated at room temperature for at least 1 hour. The filter paper was then discarded, and the plate was incubated at 37°C in 5% CO_2_ overnight. The following day, the agar plate was imaged on a ChemiDoc MP (Bio-Rad). Three raters independently graded the bacterial coverage on a 5-point scale, ranging from 0–4, with 0 being the lowest level of bacterial coverage and 4 being the highest. The raters assigned separate grades to the central area, where the wound was located, and the periwound area.

Bacterial distribution across wounds was also visualized by generating heatmaps in ImageJ, as previously described (Wallblom et al., 2024). Briefly, individual Bactogram images from all wounds within each treatment group and day were combined, and the resulting composite images were rendered with a colormap to indicate the frequency of bacterial presence at each location relative to the maximum possible frequency. The assessors were blinded to the identity of the assigned intervention for this measure.

### Identification of bacteria

To identify the major cultivable aerobic bacteria, including *S. aureus* and various commensal staphylococci, MALDI-TOFMS, associated with MALDI Biotyper software, was used for the rapid and specific identification at the species level (Dichtl et al., 2023, Dubois et al., 2010). Swab and dressing fluid samples were cultivated on a blood agar plate as described in (Lundgren et al., 2025). Briefly, the day after, six colonies were selected from each plate. Colonies were prepared on stainless steel MALDI target plates as described by the manufacturer (Bruker Daltronik GmbH). The target plate was analyzed by a Microflex LT/SH SMART MALDI-TOF mass spectrometer with flexControl v. 3.4 (Bruker Daltronik GmbH) The mass spectra were collected in linear mode over a mass range of 2 to 20 kDa. For each sample spot a spectrum of 240 summed laser shots was acquired. The spectra were analyzed using a MALDI Biotyper (MBT) Compass v. 4.1 with the MBT Compass Library Revision L (DB-9607, 2020) (Bruker Daltronik GmbH). For species-level determination a MALDI Biotyper score of 2.0 or more was required, except for *Corynebacterium,* for which an identification with a score of 2.0 or more was limited to the genus level, and for species that belonged to the *Bacillus cereus* complex, wherein the identification was limited to the complex level. Assessors were blinded to the identity of the assigned intervention for this measure.

### DNA extraction, polymerase chain reaction, sequencing, and bioinformatics

Prior to DNA extraction, skin swab samples were centrifuged (8000× g, 30 min at 4°C), and the supernatant was discarded. The pellets were lysed by using lysostaphin (0.05 mg/mL, Sigma, Burlington, VT, USA) and lysozyme (9.5 mg/mL, Sigma, Burlington, VT, USA) for 1 h. DNA was extracted using the DNeasy PowerSoil Kit (Qiagen, Hilden, Germany), following the manufacturer’s instructions. DNA concentrations were measured with the Qubit dsDNA HS Assay (ThermoFisher Scientific, Waltham, MA, USA) using a Qubit fluorometer.

The bacterial population was analyzed by amplification of the 16S rRNA gene fragment using the V1V3 primer set (fw, 5’-AGAGTTTGATCCTGGCTCAG-3’; rev, 5’-ATYACCGCGGCTGCTGGCA-3’). The *tuf2* PCR (for staphylococcal population analysis) was performed as described previously (Feidenhansl et al., 2024) using the primers tuf2_fw, 5′-ACAGGCCGTGTTGAACGTG-3′ and tuf2_rev, 5′-ACAGTACGTCCACCTTCACG-3′. PCR reaction mixtures were made in a total volume of 25 µL and comprised 5 µL of DNA sample, 2.5 µL AccuPrime PCR Buffer II (Invitrogen, Waltham, MA, USA), 1.5 µL of each primer (10 µM) (DNA Technology, Risskov, Denmark), 0.15 µL AccuPrime Taq DNA Polymerase High Fidelity (Invitrogen, Waltham, USA), and 14.35 µL of PCR grade water. The PCR reaction was performed using the following cycle conditions: initial denaturation at 94°C for 2 min, 35 cycles of denaturation at 94°C for 20 sec, annealing at 55°C for 30 sec, elongation at 68°C for 1 min, and final elongation step at 72°C for 5 min. PCR products were verified on an agarose gel and purified using the Qiagen GeneReadTM Size Selection kit (Qiagen, Hilden, Germany). The concentration of the purified PCR products was measured with a NanoDrop 2000 spectrophotometer (ThermoFisher Scientific, Waltham, MA, USA).

Specific indices and Illumina adapters were attached to the amplicons using the Nextera XT Index kit (Illumina, San Diego, CA, USA). Index PCR was performed using 5 µL of template PCR product, 2.5 µL of each index primer, 12.5 µL of 2× KAPA HiFi HotStart ReadyMix, and 2.5 µL PCR grade water. The thermal cycling scheme was as follows: 95°C for 3 min, 8 cycles of 30 s at 95°C, 30 s at 55°C, and 30 s at 72°C and a final extension at 72°C for 5 min. Quantification of the products was performed using the Quant-iT dsDNA HS assay kit and a Qubit fluorometer (Invitrogen GmbH, Karlsruhe, Germany) following the manufacturer’s instructions. MagSi-NGSPREP Plus Magnetic beads (Steinbrenner Laborsysteme GmbH, Wiesenbach, Germany) were used for purification of the indexed products as recommended by the manufacturer, and normalization was performed using the Janus Automated Workstation from Perkin Elmer (Perkin Elmer, Waltham, MA, USA). Sequencing was conducted with an Illumina MiSeq platform using dual indexing and the MiSeq reagent kit v3 (600 cycles) as recommended by the manufacturer.

Sequences (FASTQ format) obtained after demultiplexing the reads and trimming the primers (Cutadapt v. 3.7) were processed with QIIME2 (v.2023.5) (Bolyen et al., 2019). Paired-end reads were denoised with DADA2 via q2-dada2 (Callahan et al., 2016); reads with more than two expected errors in either the forward or reverse reads were discarded, and chimeras were removed.

Unique sequences obtained through DADA2 were clustered with VSEARCH (Rognes et al., 2016) using q2-vsearch (Rideout et al., 2014) at a 99% identity cut-off against allele databases. 16S rRNA sequence reads were classified using a 16S rRNA-based feature-classifier (via q2-feature-classifier) based on the V1-V3 regions of SILVA 138 99% OTUs (Pruesse et al., 2007). The database for the staphylococcal amplicon scheme contained all *tuf2* alleles from staphylococcal genomes available in GenBank (as of January 2025). Data were normalized, and low abundant features were filtered.

### Statistical analysis

A statistical analysis plan was prepared before analyses were initiated. The primary analyses of cytokines/chemokines, neutrophil proteins, wound leakage, and bacterial levels described below were conducted according to this plan. All other described analyses were post hoc. Statisticians were not blinded to the identity of the assigned intervention for any analyses.

#### Primary analysis of cytokines/chemokines and neutrophil proteins

All data are presented using box plots indicating the median, interquartile range, and range. Data were plotted using GraphPad Prism 10 software. When a logarithmic axis was used, zero values were replaced with 1, if not otherwise specified, to enable visualization of all data points on the graph but were not replaced in any statistical analyses.

#### Primary analysis of wound leakage

Data were visualized and presented using box plots as described above. Protein content, a continuous variable, was compared in TCP-25-treated and control wounds using a linear mixed effects model, with the results presented as mean differences with 95% confidence intervals and *P* values. Models were constructed with protein content as a dependent variable. Wound location (proximal or distal, left leg or right leg), treatment (TCP-25 or control gel), time (all three modelled as fixed effects) and study participants (modelled as random effects) were independent variables. Models were constructed separately for the 3 dose groups (consisting of 32 wounds each, 16 treated with TCP-25 and 16 treated with control gel). Adherence to the model conditions was tested using residual diagnostic plots. *P* values were not adjusted for multiple comparisons due to the exploratory nature of the study. The data were not transformed.

#### Primary analysis of bacterial levels

Data were visualized and presented using box plots as described above. Bacterial load in swab and dressing fluid samples (continuous variables) were compared in TCP-25-treated and control wounds using a linear mixed-effects model constructed as described above. Because bacterial levels were very low in all samples on Days 1 and 2, which violated model conditions, Days 1 and 2 were excluded from the mixed models. Because of heavily skewed data, the data were transformed so the dependent variable became the square root of the bacterial count. *P* values were not adjusted for multiple comparisons due to the exploratory nature of the study. Predicted bacterial loads were calculated on the population level for the average CFU of the two wounds on each participant that received the same treatment. A sensitivity analysis excluding the outliers was carried out as above.

#### Exploratory post hoc analyses of cytokine, neutrophil protein, protein leakage, and bacterial levels

To explore time-specific differences in statistical significance on cytokine and neutrophil protein levels, wound leakage, and bacterial burden, each parameter was compared in TCP-25-treated and control wounds at each time point using a matched-pairs Wilcoxon signed-rank test, in R (v4.5.2). Pratt’s method was applied for handling zero differences. Each participant had 2 wounds that were treated with TCP-25 gel and 2 wounds that were treated with placebo gel. To obtain independent values and mitigate within-subject effects, the average of the values of the two wounds from each participant that received the same treatment (TCP-25 or placebo) was calculated and used in the analysis, resulting in 8 values per group for each analysis. Because the test calculates the sum of the ranks of the differences and because there were only 8 values per group in each analysis, the lowest *P* value achievable in this analysis is 0.0078 and indicates that the level was higher in the control group in every instance. *P* values were not adjusted for multiplicity as this was an exploratory analysis intended to generate a visualization of effects and not intended as confirmatory significance testing. *P* values < 0.05 were considered statistically significant.

#### Microbiome composition and diversity

Statistical analyses of microbiome data were performed in R (v4.5.2). Non-bacterial reads (e.g., chloroplast) were removed, OTUs below 1% relative abundance per sample were collapsed as "under threshold," and remaining abundances were renormalized. Sample sizes per analysis are given in the figure and table legends; full relative-abundance profiles (16S V1–V3, genus level; *tuf2*, *Staphylococcus* species level) are in Supplementary data.

Alpha diversity (richness, Shannon index, Pielou’s evenness; vegan v2.7-2) was modeled with dose-stratified linear mixed-effects models, as described above. Richness was log(1+x)-transformed and Shannon and Pielou square-root-transformed to meet model assumptions; samples with richness ≤ 1 (undefined Pielou) were excluded. Marginal means were back-transformed (emmeans v2.0.0), and variance explained by treatment (partial R²) and by all fixed effects (conditional R²) was estimated with partR2 v0.9.2.

Beta diversity was assessed from Bray–Curtis distances (vegan), visualized by PCoA (wcmdscale), and tested with dose-stratified PERMANOVA (adonis2, 9999 permutations) on Treatment and Timepoint, with permutations constrained within Participant (strata). The Treatment × Timepoint interaction was tested first; if non-significant, the Treatment main effect was estimated adjusting for Timepoint. Group dispersions were compared with PERMDISP (betadisper).

### Study approval

The trial and collection of specimens to the biobank was approved by the Swedish ethical review authority (etikprövningsmyndigheten application number 2022-00527-01). Written informed consent was received from all subjects prior to participation.

## Data Availability

All data produced in the present study are available upon reasonable request to the authors

## Author contributions

The following authors contributed to each of the following roles (defined according to the CRediT taxonomy): Conceptualization, GP, KW, SL, KS, MP, AS; design of methodology, GP, KW, SL, MP and AS; formal analysis (statistics), GP, KW, JFPC, CL, JF; investigation (performing experiments and data collection) and validation, GP, KW, BN, JFPC, ACS, FF, CL, HB; resources (provision of participants and samples), KW, SL, KS, AS; data curation, GP, KW, JFPC, JF; writing (original draft preparation), GP, KW, JF, AS; writing (review and editing), GP, KW, SL, BN, JFPC, ACS, FF, CL, JF, KS, MP, HB, AS.; visualization, GP, KW, JFPC, FF, CL, EH, JF; supervision, GP, KS, AS; project administration, GP, KW, JF, AS; funding acquisition, AS. All authors have read and agreed to the published version of the manuscript.

## Acknowledgments

The exploratory data presented here was supported by grants from the Swedish Research Council (2020-02016, 2025-02401), Edvard Welanders Stiftelse and Finsenstiftelsen (Hudfonden), the Royal Physiographic Society, the Crafoord and Österlund Foundations, and the Swedish Government Funds for Clinical Research (ALF). Xinnate AB provided the project management resources and expertise for the regulatory development enabling the clinical parts of the Safety study that generated the control samples used in this work. We thank Susanne Erdmann and Anne Nielsen and other personnel at the Department of Dermatology Lund and the Clinical Trial Unit at Skane University Hospital Lund for support. We thank Sean Kim at AdvanSci Research Solutions and Blue Pencil Science for support with medical writing.

## Conflict of interest disclosure statement

AS is a founder of in2cure AB, a parent company of Xinnate AB which was the sponsor of the clinical trial from which the biobank samples used in this study are derived. GP was employed part-time (20%) by Xinnate AB during the study. JF provided consulting services to Xinnate AB during the study. The other authors have declared that no conflict of interest exists.

## Notes

### Clinical Trial

NCT05378997

### Author Declarations

Ethics committee/IRB of the Swedish Ethical Review Authority (Etikprövningsmyndigheten) gave ethical approval for this work. Approval number 2022-00527-01.

## References

Beutler BA. TLRs and innate immunity. Blood 2009;113(7):1399–407.

Bolyen E, Rideout JR, Dillon MR, Bokulich NA, Abnet CC, Al-Ghalith GA, et al. Reproducible, interactive, scalable and extensible microbiome data science using QIIME 2. Nat Biotechnol 2019;37(8):852–7.

Callahan BJ, McMurdie PJ, Rosen MJ, Han AW, Johnson AJA, Holmes SP. DADA2: High-resolution sample inference from Illumina amplicon data. Nature Methods 2016;13(7):581–3.

Canchy L, Kerob D, Demessant A, Amici JM. Wound healing and microbiome, an unexpected relationship. J Eur Acad Dermatol Venereol 2023;37 Suppl 3:7–15.

Dichtl K, Klugherz I, Greimel H, Luxner J, Koberl J, Friedl S, et al. A head-to-head comparison of three MALDI-TOF mass spectrometry systems with 16S rRNA gene sequencing. J Clin Microbiol 2023;61(10):e0191322.

Dubois D, Leyssene D, Chacornac JP, Kostrzewa M, Schmit PO, Talon R, et al. Identification of a variety of Staphylococcus species by matrix-assisted laser desorption ionization-time of flight mass spectrometry. J Clin Microbiol 2010;48(3):941–5.

Ellis S, Lin EJ, Tartar D. Immunology of Wound Healing. Curr Dermatol Rep 2018;7(4):350–8.

Eming SA, Krieg T, Davidson JM. Inflammation in wound repair: molecular and cellular mechanisms. J Invest Dermatol 2007;127(3):514–25.

Feidenhansl C, Lund M, Poehlein A, Lood R, Lomholt HB, Brüggemann H. Cutibacterium and Staphylococcus dysbiosis of the skin microbiome in acne and its decline after isotretinoin treatment. JEADV Clinical Practice 2024;3(5):1454–66.

Gurtner GC, Werner S, Barrandon Y, Longaker MT. Wound repair and regeneration. Nature 2008;453(7193):314–21.

Hansen FC, Kalle-Brune M, van der Plas MJ, Stromdahl AC, Malmsten M, Morgelin M, et al. The Thrombin-Derived Host Defense Peptide GKY25 Inhibits Endotoxin-Induced Responses through Interactions with Lipopolysaccharide and Macrophages/Monocytes. J Immunol 2015;194(11):5397–406.

Lim CH, Puthia M, Butrym M, Tay HM, Lee MZY, Hou HW, et al. Thrombin-derived host defence peptide modulates neutrophil rolling and migration in vitro and functional response in vivo. Scientific Reports 2017;7(1):11201.

Lundgren S, Petruk G, Wallblom K, Cardoso JFP, Stromdahl AC, Forsberg F, et al. Temporal dynamics and interrelations of cytokines, neutrophil proteins, exudation, and bacterial colonization in epidermal wound healing. Front Med (Lausanne) 2025;12:1609347.

Lundgren S, Wallblom K, Fisher J, Erdmann S, Schmidtchen A, Saleh K. Study protocol for a phase 1, randomised, double-blind, placebo-controlled study to investigate the safety, tolerability and pharmacokinetics of ascending topical doses of TCP-25 applied to epidermal suction blister wounds in healthy male and female volunteers. BMJ Open 2023;13(2):e064866.

Lundqvist K, Herwald H, Sonesson A, Schmidtchen A. Heparin binding protein is increased in chronic leg ulcer fluid and released from granulocytes by secreted products of Pseudomonas aeruginosa. Thromb Haemost 2004;92(2):281–7.

Odobasic D, Kitching AR, Yang Y, O’Sullivan KM, Muljadi RC, Edgtton KL, et al. Neutrophil myeloperoxidase regulates T-cell-driven tissue inflammation in mice by inhibiting dendritic cell function. Blood 2013;121(20):4195–204.

Papareddy P, Rydengard V, Pasupuleti M, Walse B, Morgelin M, Chalupka A, et al. Proteolysis of human thrombin generates novel host defense peptides. PLoS Pathog 2010;6(4):e1000857.

Prat M, Guenezan J, Drugeon B, Burucoa C, Mimoz O, Pichon M. Impact of Skin Disinfection on Cutaneous Microbiota, before and after Peripheral Venous Catheter Insertion. Antibiotics (Basel) 2022;11(9).

Pruesse E, Quast C, Knittel K, Fuchs BM, Ludwig W, Peplies J, et al. SILVA: a comprehensive online resource for quality checked and aligned ribosomal RNA sequence data compatible with ARB. Nucleic Acids Research 2007;35(21):7188–96.

Puthia M, Butrym M, Petrlova J, Stromdahl AC, Andersson MA, Kjellstrom S, et al. A dual-action peptide-containing hydrogel targets wound infection and inflammation. Sci Transl Med 2020;12(524).

Rideout JR, He Y, Navas-Molina JA, Walters WA, Ursell LK, Gibbons SM, et al. Subsampled open-reference clustering creates consistent, comprehensive OTU definitions and scales to billions of sequences. PeerJ 2014;2:e545.

Rognes T, Flouri T, Nichols B, Quince C, Mahe F. VSEARCH: a versatile open source tool for metagenomics. PeerJ 2016;4:e2584.

Saleh K, Sonesson A, Persson K, Riesbeck K, Schmidtchen A. Can dressings soaked with polyhexanide reduce bacterial loads in full-thickness skin grafting? A randomized controlled trial. J Am Acad Dermatol 2016;75(6):1221–8.e4.

SanMiguel AJ, Meisel JS, Horwinski J, Zheng Q, Bradley CW, Grice EA. Antiseptic Agents Elicit Short-Term, Personalized, and Body Site-Specific Shifts in Resident Skin Bacterial Communities. J Invest Dermatol 2018;138(10):2234–43.

Saravanan R, Adav SS, Choong YK, van der Plas MJA, Petrlova J, Kjellstrom S, et al. Proteolytic signatures define unique thrombin-derived peptides present in human wound fluid in vivo. Sci Rep 2017;7(1):13136.

Saravanan R, Holdbrook DA, Petrlova J, Singh S, Berglund NA, Choong YK, et al. Structural basis for endotoxin neutralisation and anti-inflammatory activity of thrombin-derived C-terminal peptides. Nat Commun 2018;9(1):2762.

Soehnlein O, Lindbom L. Neutrophil-derived azurocidin alarms the immune system. J Leukoc Biol 2009;85(3):344–51.

Storm-Versloot MN, Vos CG, Ubbink DT, Vermeulen H. Topical silver for preventing wound infection. Cochrane Database Syst Rev 2010(3):Cd006478.

Stromdahl AC, Ignatowicz L, Petruk G, Butrym M, Wasserstrom S, Schmidtchen A, et al. Peptide-coated polyurethane material reduces wound infection and inflammation. Acta Biomater 2021.

Takeuchi O, Akira S. Pattern recognition receptors and inflammation. Cell 2010;140(6):805–20.

Tomic-Canic M, Burgess JL, O’Neill KE, Strbo N, Pastar I. Skin Microbiota and its Interplay with Wound Healing. Am J Clin Dermatol 2020;21(Suppl 1):36–43.

Walker A, Brace J. A multipurpose dressing: role of a Hydrofiber foam dressing in managing wound exudate. J Wound Care 2019;28(Sup9a):S4–s10.

Wallblom K, Forsberg F, Lundgren S, Fisher J, Cardoso J, Petruk G, et al. Bactogram: Spatial Analysis of Bacterial Colonisation in Epidermal Wounds. Exp Dermatol 2024;33(12):e70018.

Wallblom K, Holmgren K, Lundgren S, Belfrage E, Hoppe T, Hugerth M, et al. Results of a non-randomized, open-label phase I study evaluating the novel immunomodulatory peptide TCP-25 for treatment of dystrophic epidermolysis bullosa. Orphanet J Rare Dis 2025;21(1):4.

Wallblom K, Lundgren S, Petruk G, Puthia M, Fisher J, Hugerth M, et al. A First-In-Human Randomized Controlled Phase 1 Study Assessing the Safety and Tolerability of Topical TCP-25 Gel in Epidermal Suction Blister Wounds. Clin Transl Sci 2026;19(2):e70497.

Young RE, Thompson RD, Larbi KY, La M, Roberts CE, Shapiro SD, et al. Neutrophil Elastase (NE)-Deficient Mice Demonstrate a Nonredundant Role for NE in Neutrophil Migration, Generation of Proinflammatory Mediators, and Phagocytosis in Response to Zymosan Particles In Vivo1. The Journal of Immunology 2004;172(7):4493–502.

Zheng Y, Hunt RL, Villaruz AE, Fisher EL, Liu R, Liu Q, et al. Commensal Staphylococcus epidermidis contributes to skin barrier homeostasis by generating protective ceramides. Cell Host Microbe 2022;30(3):301–13 e9.

